# Assessing the Potential of AI-Driven Drug Repurposing in Ophthalmology: An Analysis of ChatGPT’s Therapeutic Recommendations

**DOI:** 10.1101/2025.06.09.25329271

**Authors:** Raziyeh Mahmoudzadeh, Michelle Zaichik, Kavin Selvan, Taharah Islam, Mirataollah Salabati, Christopher T. Leffler

## Abstract

**Purpose:** This study aimed to evaluate the novelty and potential value of therapeutic suggestions made by an artificial intelligence large language model for treating various ophthalmic diseases.

**Methods:** ChatGPT-3.5 was used to suggest novel ophthalmic indications for available medications. The generation of therapeutic suggestions was performed by inputting standardized queries about treatments for common ophthalmic conditions and then categorizing the responses by drug type. Data tables were organized by ophthalmic condition, with consistent quality checks to ensure accuracy. Literature searches were conducted to determine the FDA-approval status of each therapy, and whether the suggested application was novel in the context of the condition. Therapies were categorized according to current use and level of evidence for use.

**Results:** ChatGPT proposed 180 medications and treatment options for 36 eye conditions. Of the 180 medications, 143 (79.4%) were FDA-approved for general medical use and 32 out of 180 (17.7%) were specifically approved for the recommended ophthalmological conditions. The majority of suggested treatments were for corneal and anterior segment disease (82/180 or 46%), with other categories being retina (23%), glaucoma (8.9%), pediatrics and strabismus (12%), neuro-ophthalmology (0.55%), and uveitis (10%). The proposed treatments were then evaluated by the degree to which the literature supported additional investigation. The majority, 86/180 (48%), were already being used in the clinic, while 27/180 (15%) represented a novel ophthalmic use that appeared to be a reasonable hypothesis to test, and 20/180 (11%) were novel, but appeared unlikely to succeed, based on their mechanism of action. The level of novelty for each treatment was also evaluated, with categories spanning from pre-existing testing in animal models to repurposed for novel ophthalmic use.

**Conclusion:** These findings suggest that ChatGPT is capable of formulating novel treatment options for a range of ophthalmic diseases. Of the suggestions, 27/180 (15%) appeared novel, and reasonable suggestions, based on their mechanism of action. ChatGPT can potentially suggest novel ophthalmic applications for existing medications, which could be evaluated with further laboratory and clinical research.

## Introduction

ChatGPT and other forms of artificial intelligence are increasingly being used to generate new ideas, form healthcare plans, and even provide novel solutions. Although ChatGPT is a relatively recent innovation, it has already undergone several updates, and its usage is becoming increasingly widespread. It has been used in medicine and is known to go beyond typical knowledge to combine what is known and unknown into the formation of new suggestions [1].

ChatGPT has been used to generate novel ideas in various fields of medicine, with more research continuously being conducted. The US Food and Drug Administration (FDA) has recently implemented a category termed Software as Medical Device (SaMD) for the use of artificial intelligence in medical treatments, with analysis done to determine patient safety and benefits before allowing for AI-based medical device approval [2]. Similarly, researchers Wang, Feng & Wei created a mathematical AI model termed SGNC to generate novel molecules that would potentially serve as effective treatments for individuals with cocaine addiction [3]. ChatGPT has also demonstrated the capability to understand proper treatments in the field of neurology, as ChatGPT-4 was found to have a 75% accuracy rate in the treatment plans for various brain tumor cases, as determined by 20 independent neurosurgeons [4]. Outside of medicine, ChatGPT generated highly novel concepts in product design innovation [5]. The study utilized a novelty metric that spanned from known, partially novel, and fully known to analyze the novelty of ChatGPT ideas.

The main modalities of artificial intelligence in ophthalmology to date have been regarding triage and diagnosis of disease, qualification of ophthalmic diseases, and answering specific questions regarding these diseases [6]. Moreover, Ming et al predicted that the capabilities of a large language model (LLM) like ChatGPT, will allow for researchers to identify novel drug targets in uveitis treatment, allowing for a more streamlined innovation of medications [7]. However, the study did not evaluate the ability of current artificial intelligence models, such as ChatGPT, to generate novel drug uses. Of significance, researchers Huang et al found that LLM chatbots can match retina specialists and outperform glaucoma specialists in the accuracy of diagnosis and treatment suggestions, highlighting the potential as valuable tools in ophthalmology [8].

Our study aims to investigate the novelty of drug suggestions by ChatGPT in the treatment of various ophthalmic diseases. Some medications have been used in ophthalmology and other fields of medicine “off-label”, for purposes outside the original approved indication. However, this paper further explores if these known medications have the possibility for use in the treatment of ophthalmic diseases as recommended by ChatGPT. While there are many emerging studies on the quality and accuracy of ChatGPT, there is little data on its performance in novel care suggestions specific to ophthalmology.

## Methods

### Study Design

This study is designed to evaluate the novelty of therapeutic suggestions made by ChatGPT for the treatment of various ophthalmic diseases. ChatGPT output was verified through a literature search of the therapies in Google Scholar. The study was approved by the Office of Research Subjects Protection of Virginia Commonwealth University.

### ChatGPT Model

The version of ChatGPT used in this study is the January 2024, ChatGPT-3.5, based on knowledge up to January 2022. ChatGPT-3.5 is a large language model (LLM) developed by OpenAI (San Francisco, US), designed to generate human-like text based on the input it receives. The model was accessed through the OpenAI platform and used its API for generating responses to specific medical queries related to ophthalmology.

## Data Collection

### Therapeutic Suggestion Generation

We inputted multiple standardized queries related to the treatment of various ophthalmic diseases, including conditions such as uveitis, glaucoma, and retinal diseases, into ChatGPT-3.5. ChatGPT was then asked to elaborate on treatments for each condition until it either exhausted its responses or began repeating them, at which point the query was changed to the next condition. These queries were designed based on common ophthalmic conditions. The responses generated by ChatGPT were recorded and initially categorized based on the type of drug suggested. We then reviewed the raw output to identify all therapies and conditions mentioned by ChatGPT. The information was first organized in a table by therapies and later restructured to list drugs by condition. Next, we performed quality checks on both tables, ensuring they accurately reflected the transcripts.

### Literature Search and Data Collection

The list of therapies was distributed among the coauthors to ensure consistent literature reviews, with each therapy being evaluated by at least two coauthors. We performed literature searches to verify whether each therapy was FDA-approved in general, utilizing the FDA’s Drug Approval Database [9] and whether they were already FDA-approved for the specific ophthalmological condition in question. We also assessed the novelty of the therapies in the context of the condition. Each therapy was categorized by its current use and evidence level: 1) Clinical application (already used for this indication) (C); 2) Cell-based/bench studies (B); 3) Animal model studies (A); 4) repurposed for a novel ophthalmic use (N); and 5) novel but supported by existing, limited literature (L); 6) novel, not supported by literature review and unlikely to be a productive line of investigation (E). For novel therapies, potential mechanisms of action were also explored. The suggested ophthalmic diseases were organized into the following primary categories: Retina, Cornea and Anterior Segment, Glaucoma, Pediatrics and Strabismus, Neuro-Ophthalmology, and Uveitis. Finally, we performed quality checks to ensure that the final table accurately reflected the research findings and discussions.

Summary statistics were calculated for FDA approval and novelty in ophthalmic conditions, using Microsoft Excel 365.

## Results

ChatGPT proposed 180 medications and treatment methods for ophthalmology. Each medication was recommended for one or more specific eye conditions and a total of 36 eye conditions were suggested.

For instance, antioxidants were suggested for treating conditions such as wet and dry age-related macular degeneration (AMD), cataracts, and diabetic retinopathy. Initially, the ocular conditions were grouped into 57 categories. However, 21 of these categories were broad and non-specific, often overlapping with more specific conditions already listed. These broad categories were excluded before conducting the literature review. For instance, categories like ocular surface disorders influenced by hormonal changes, and lacrimal gland syndromes were removed as they were too broad or already addressed in more specific categories like dry eye disease. In this paper, onchocerciasis was classified as a corneal disease. Ultimately, 36 eye conditions were included as shown in Table 1.

**Table 1.** Potential Novel Therapeutic Suggestions by ChatGPT Including all Ophthalmic Categories.

| Disease | Drug(s) | FDA-approval status | Current status of drug for this indication |
| --- | --- | --- | --- |
| age-related macular degeneration | antioxidants and nutritional supplements | Available without FDA approval | Already used for this indication |
| age-related macular degeneration | lutein and zeaxanthin | Not FDA-approved | Already used for this indication |
| age-related macular degeneration | metformin | FDA-approved for other indications | Not used routinely, supported by limited literature in humans |
| age-related macular degeneration | methylene blue | FDA-approved for other indications | Animal model studies |
| age-related macular degeneration | omega-Glaucoma fatty acids | Available without FDA approval | Already used for this indication |
| age-related macular degeneration | rapamycin (sirolimus) | Not FDA-approved | Animal model studies |
| age-related macular degeneration | resveratrol | Not FDA-approved | Animal model studies |
| age-related macular degeneration | taurine | Not FDA-approved | Animal model studies |
| allergic conjunctivitis | azelastine | Already FDA-approved for this indication | Already used for this indication |
| allergic conjunctivitis | montelukast | FDA-approved for other indications | Not used routinely, supported by limited literature in humans |
| allergic conjunctivitis | olopatadine | Already FDA-approved for this indication | Already used for this indication |
| amblyopia | antioxidants | Not FDA-approved | Novel (used in humans for other purposes) |
| amblyopia | atomoxetine | Not FDA-approved | Novel, however not supported by literature and unlikely to succeed |
| amblyopia | binocular vision therapy | NA | Already used for this indication |
| amblyopia | cycloplegic agents (e.g., atropine) | FDA-approved for other indications | Already used for this indication |
| amblyopia | levetiracetam | FDA-approved for other indications | Novel, however not supported by literature and unlikely to succeed |
| amblyopia | lurasidone | FDA-approved for other indications | Novel, however not supported by literature and unlikely to succeed |
| amblyopia | lutein and zeaxanthin | Available without FDA approval | Novel (used in human for other purposes) |
| amblyopia | memantine | FDA-approved for other indications | Animal model studies |
| amblyopia | omega-3 fatty acids | Available without FDA approval | Not used routinely, supported by limited literature in humans |
| amblyopia | perceptual learning | Not FDA-approved | Already used for this indication |
| amblyopia | rivastigmine | FDA-approved for other indications | Novel, however not supported by literature and unlikely to succeed |
| amblyopia | transcranial magnetic stimulation (TMS) | FDA-approved for other indications | Novel (used in human for other purposes) |
| amblyopia | virtual reality (VR) therapy | Not FDA-approved | Already used for this indication |
| astigmatism | cycloplegic agents (e.g., atropine) | FDA-approved for other indications | Novel (used in human for other purposes) |
| astigmatism | omega-3 fatty acids | Available without FDA approval | Novel (used in human for other purposes) |
| astigmatism | riboflavin (vitamin B2) | Available without FDA approval | Already used for this indication |
| astigmatism | vitamin C (ascorbic acid) | Available without FDA approval | Not used routinely, supported by limited literature in humans |
| bacterial conjunctivitis | erythromycin | Already FDA-approved for this indication | Already used for this indication |
| bacterial conjunctivitis | moxifloxacin | Already FDA-approved for this indication | Already used for this indication |
| blepharitis | azithromycin | FDA-approved for other indications | Already used for this indication |
| blepharitis | brimonidine tartrate | FDA-approved for other indications | Not used routinely, supported by limited literature in humans |
| blepharitis | cyclosporine | FDA-approved for other indications | Already used for this indication |
| blepharitis | doxycycline | FDA-approved for other indications | Already used for this indication |
| blepharitis | macrocyclic lactones (e.g., ivermectin) | FDA-approved for other indications | Not used routinely, supported by limited literature in humans |
| blepharitis | minocycline | FDA-approved for other indications | Already used for this indication |
| blepharitis | tacrolimus | FDA-approved for other indications | Not used routinely, supported by limited literature in humans |
| cataracts | angiotensin-converting enzyme (ACE) inhibitors (e.g., lisinopril, enalapril) | FDA-approved for other indications | Not used routinely, supported by limited literature in humans |
| cataracts | aspirin (acetylsalicylic acid) | FDA-approved for other indications | Not used routinely, supported by limited literature in humans |
| cataracts | atorvastatin | FDA-approved for other indications | Not used routinely, supported by limited literature in humans |
| cataracts | N-acetylcarnosine | Not FDA-approved | Not used routinely, supported by limited literature in humans |
| cataracts | pirenoxine (cataractin) | Not FDA-approved | Already used for this indication |
| certain forms of macular degeneration | pentoxifylline | FDA-approved for other indications | Not used routinely, supported by limited literature in humans |
| certain forms of retinopathy | dipyridamole | FDA-approved for other indications | Novel (used in human for other purposes) |
| optic neuropathy | pioglitazone | FDA-approved for other indications | Animal model studies |
| conjunctivitis | acitretin | FDA-approved for other indications | Novel, however not supported by literature and unlikely to succeed |
| corneal ulcers | amphotericin B | FDA-approved for other indications | Already used for this indication |
| corneal ulcers | ciprofloxacin/ofloxacin | Already FDA-approved for this indication | Already used for this indication |
| corneal ulcers | cyclosporine | FDA-approved for other indications | Novel (used in human for other purposes) |
| corneal ulcers | doxycycline | FDA-approved for other indications | Already used for this indication |
| corneal ulcers | epidermal growth factor (EGF) eye drops | Not FDA-approved | Already used for this indication |
| corneal ulcers | natamycin | Already FDA-approved for this indication | Already used for this indication |
| corneal ulcers | tacrolimus | FDA-approved for other indications | Already used for this indication |
| corneal ulcers | tobramycin | Already FDA-approved for this indication | Already used for this indication |
| demodex infestation | macrocyclic lactones (e.g., ivermectin) | FDA-approved for other indications | Already used for this indication |
| diabetic retinopathy | angiotensin-converting enzyme (ACE) inhibitors (e.g., lisinopril, enalapril) | FDA-approved for other indications | Already used for this indication |
| diabetic retinopathy | anti-VEGF agents (e.g. ranibizumab) | Already FDA-approved for this indication | Already used for this indication |
| diabetic retinopathy | atorvastatin | FDA-approved for other indications | Novel (used in human for other purposes) |
| diabetic retinopathy | fenofibrate | FDA-approved for other indications | Novel (used in human for other purposes) |
| diabetic retinopathy | metformin | FDA-approved for other indications | Not used routinely, supported by limited literature in humans |
| diabetic retinopathy | omega-3 fatty acids or supplements | Available without FDA approval | Already used for this indication |
| diabetic retinopathy | pentoxifylline | FDA-approved for other indications | Not used routinely, supported by limited literature in humans |
| diabetic retinopathy | pioglitazone | FDA-approved for other indications | Already used for this indication |
| diabetic retinopathy | ranibizumab/aflibercept | Already FDA-approved for this indication | Already used for this indication |
| diabetic retinopathy | taurine | Not FDA-approved | Not used routinely, supported by limited literature in humans |
| dry age-related macular degeneration | antioxidants and nutritional supplements | Available without FDA approval. | Not used routinely, supported by limited literature in humans |
| dry age-related macular degeneration | curcumin | Available without FDA approval. | Not used routinely, supported by limited literature in humans |
| dry age-related macular degeneration | DHA (docosahexaenoic acid) | Not FDA-approved | Not used routinely, supported by limited literature in humans |
| dry age-related macular degeneration | fenofibrate | FDA-approved for other indications | Not used routinely, supported by limited literature in humans |
| dry age-related macular degeneration | memantine | FDA-approved for other indications | Cell-based/bench studies |
| dry age-related macular degeneration | omega-3 fatty acids or supplements | Available without FDA approval | Already used for this indication |
| dry age-related macular degeneration | rapamycin (sirolimus) | FDA-approved for other indications | Novel (used in human for other purposes) |
| dry age-related macular degeneration | taurine | Available without FDA approval | Animal model studies |
| dry eye syndrome | artificial tear supplements | Already FDA-approved for this indication | Already used for this indication |
| dry eye syndrome | autologous serum tears | Not FDA-approved | Already used for this indication |
| dry eye syndrome | cyclosporine | Already FDA-approved for this indication | Already used for this indication |
| dry eye syndrome | diquafosol | Not FDA-approved | Already used for this indication |
| dry eye syndrome | lifitegrast | Already FDA-approved for this indication | Already used for this indication |
| dry eye syndrome | N-acetylcysteine (NAC) | FDA-approved for other indications | Already used for this indication |
| dry eye syndrome | tacrolimus | FDA-approved for other indications | Novel (used in human for other purposes) |
| dry eye syndrome | tamoxifen | FDA-approved for other indications | Novel, however not supported by literature and unlikely to succeed |
| eye disorders related to compromised blood flow | pentoxifylline | FDA-approved for other indications | Novel (used in human for other purposes) |
| glaucoma | apraclonidine | Already FDA-approved for this indication | Already used for this indication |
| glaucoma | betaxolol or other beta-blockers | Already FDA-approved for this indication | Already used for this indication |
| glaucoma | brimonidine tartrate | Already FDA-approved for this indication | Already used for this indication |
| glaucoma | calcium channel blockers (e.g., amlodipine) | FDA-approved for other indications | Novel (used in human for other purposes) |
| glaucoma | carbonic anhydrase inhibitors | Already FDA-approved for this indication | Already used for this indication |
| glaucoma | dipivefrin | Not FDA-approved | Already used for this indication |
| glaucoma | dipyridamole | FDA-approved for other indications | Novel, however not supported by literature and unlikely to succeed |
| glaucoma | dorzolamide/timolol combination | Already FDA-approved for this indication | Already used for this indication |
| glaucoma | latanoprost | Already FDA-approved for this indication | Already used for this indication |
| glaucoma | melatonin | Not FDA-approved | Novel (used in human for other purposes) |
| glaucoma | minoxidil | FDA-approved for other indications | Not used routinely, supported by limited literature in humans |
| glaucoma | nicergoline | Not FDA-approved | Novel (used in human for other purposes) |
| glaucoma | nitric oxide donors (e.g., nitroglycerin) | FDA-approved for other indications | Novel (used in human for other purposes) |
| glaucoma | pilocarpine | FDA-approved for other indications | Already used for this indication |
| glaucoma | rho kinase inhibitors (e.g., ripasudil) | Not FDA-approved | Already used for this indication |
| glaucoma | timolol (as in "timoptic") | Already FDA-approved for this indication | Already used for this indication |
| hyperopia | brimonidine tartrate | FDA-approved for other indications | Animal model studies |
| hyperopia | cyclopentolate | FDA-approved for other indications | Already used for this indication |
| hyperopia | cycloplegic agents (e.g., atropine) | FDA-approved for other indications | Already used for this indication |
| hyperopia | omega-3 fatty acids or supplements | Available without FDA approval | Novel (used in human for other purposes) |
| hyperopia | pilocarpine | FDA-approved for other indications | Novel (used in human for other purposes) |
| inflammatory eye diseases | thalidomide | FDA-approved for other indications | Novel (used in human for other purposes) |
| meibomian gland dysfunction | N-acetylcysteine (NAC) | FDA-approved for other indications | Novel (used in human for other purposes) |
| myopia | brimonidine tartrate | FDA-approved for other indications | Novel (used in human for other purposes) |
| myopia | cyclopentolate | FDA-approved for other indications | Novel (used in human for other purposes) |
| myopia | cycloplegic agents (e.g., atropine) | FDA-approved for other indications | Already used for this indication |
| myopia | omega-3 fatty acids or supplements | Available without FDA approval | Not used routinely, supported by limited literature in humans |
| myopia | pilocarpine | FDA-approved for other indications | Novel (used in human for other purposes) |
| ocular surface infections caused by parasites (Loiasis) | ivermectin | Already FDA-approved for this indication | Already used for this indication |
| traumatic hyphema | topical anesthetics (e.g. tetracaine, proparacaine) | FDA-approved for other indications | Novel, however not supported by literature and unlikely to succeed |
| traumatic hyphema | topical cycloplegics | FDA-approved for other indications | Not used routinely, supported by limited literature in humans |
| traumatic hyphema | topical steroids | FDA-approved for other indications | Not used routinely, supported by limited literature in humans |
| traumatic hyphema | tranexamic acid | FDA-approved for other indications | Not used routinely, supported by limited literature in humans |
| onchocerciasis (BUveitis Glaucoma) | macrocyclic lactones (e.g., ivermectin) | Already FDA-approved for this indication | Already used for this indication |
| onchocerciasis | azithromycin | FDA-approved for other indications | Not used routinely, supported by limited literature in humans |
| onchocerciasis | doxycycline | FDA-approved for other indications | Not used routinely, supported by limited literature in humans |
| onchocerciasis | minocycline | FDA-approved for other indications | Animal model studies |
| onchocerciasis | moxidectin | Already FDA-approved for this indication | Already used for this indication |
| onchocerciasis | rifampicin | FDA-approved for other indications | Novel (used in humans for other purposes) |
| presbyopia | carbachol | FDA-approved for other indications | Novel, however not supported by literature and unlikely to succeed |
| presbyopia | cycloplegic agents (e.g., atropine) | FDA-approved for other indications | Novel, however not supported by literature and unlikely to succeed |
| presbyopia | dorzolamide/timolol combination | FDA-approved for other indications | Novel, however not supported by literature and unlikely to succeed |
| presbyopia | latanoprostene bunod | FDA-approved for other indications | Novel, however not supported by literature and unlikely to succeed |
| presbyopia | Omega-3 fatty acids or supplements | Available without FDA approval | Already used for this indication |
| presbyopia | pilocarpine | Already FDA-approved for this indication | Already used for this indication |
| prevention of retinal detachments | fibrinolytic agents (e.g., alteplase) | FDA-approved for other indications | Not used routinely, supported by limited literature in humans |
| prevention of retinal detachments | Omega-3 fatty acids | Available without FDA approval | Already used for this indication |
| prevention of retinal detachments | pentoxifylline | FDA-approved for other indications | Novel, however not supported by literature and unlikely to succeed |
| prevention of retinal detachments | tranexamic acid | FDA-approved for other indications | Novel, however not supported by literature and unlikely to succeed |
| prevention of retinal detachments | minocycline | FDA-approved for other indications | Novel, however not supported by literature and unlikely to succeed |
| retinal detachments (treatment) | fibrinolytic agents (e.g., alteplase) | FDA-approved for other indications | Animal model studies |
| retinal detachments (treatment) | lithium | FDA-approved for other indications | Animal model studies |
| retinal detachments (treatment) | pentoxifylline | FDA-approved for other indications | Novel, however not supported by literature and unlikely to succeed |
| retinal detachments (treatment) | tranexamic acid | FDA-approved for other indications | Novel, however not supported by literature and unlikely to succeed |
| retinal hemorrhage | tranexamic acid | FDA-approved for other indications | Animal model studies |
| retinal ischemia | prazosin | FDA-approved for other indications | Animal model studies |
| retinal ischemia | sildenafil | FDA-approved for other indications | Novel (used in human for other purposes) |
| strabismus | acetazolamide | FDA-approved for other indications | Novel, however not supported by literature and unlikely to succeed |
| retinal ischemia | baclofen | FDA-approved for other indications | Novel, however not supported by literature and unlikely to succeed |
| retinal ischemia | botulinum toxin (botox) | Already FDA-approved for this indication | Already used for this indication |
| retinal ischemia | cycloplegic agents (e.g., atropine) | FDA-approved for other indications | Already used for this indication |
| retinal ischemia | dalfampridine | FDA-approved for other indications | Novel, however not supported by literature and unlikely to succeed |
| retinal ischemia | tizanidine | FDA-approved for other indications | Novel (used in human for other purposes) |
| retinal ischemia | visual stimulation methods | Not applicable | Already used for this indication |
| trachoma | azithromycin | Already FDA-approved for this indication | Already used for this indication |
| trachoma | doxycycline | Already FDA-approved for this indication | Already used for this indication |
| trachoma | erythromycin | Already FDA-approved for this indication | Already used for this indication |
| trachoma | rifampicin | FDA-approved for other indications | Already used for this indication |
| trachoma | tetracycline | Already FDA-approved for this indication | Already used for this indication |
| trachoma | topical povidone-iodine | FDA-approved for other indications | Novel (used in human for other purposes) |
| uveitis | adalimumab | Already FDA-approved for this indication | Already used for this indication |
| uveitis | infliximab | FDA-approved for other indications | Already used for this indication |
| uveitis | colchicine | FDA-approved for other indications | Not used routinely, supported by limited literature in humans |
| uveitis | cyclophosphamide | FDA-approved for other indications | Already used for this indication |
| uveitis | cyclosporine | FDA-approved for other indications | Already used for this indication |
| uveitis | dapsone | FDA-approved for other indications | Animal model studies |
| uveitis | etanercept | FDA-approved for other indications | Already used for this indication |
| uveitis | hydroxychloroquine | FDA-approved for other indications | Already used for this indication |
| uveitis | leflunomide | FDA-approved for other indications | Already used for this indication |
| uveitis | methotrexate | FDA-approved for other indications | Already used for this indication |
| uveitis | methylprednisolone | Already FDA-approved for this indication | Already used for this indication |
| uveitis | minocycline | FDA-approved for other indications | Animal model studies |
| uveitis | mycophenolate mofetil | FDA-approved for other indications | Already used for this indication |
| uveitis | rituximab | FDA-approved for other indications | Already used for this indication |
| uveitis | tacrolimus | FDA-approved for other indications | Already used for this indication |
| uveitis | thalidomide | FDA-approved for other indications | Not used routinely, supported by limited literature in humans |
| uveitis | tocilizumab | FDA-approved for other indications | Not used routinely, supported by limited literature in humans |
| vascular-related eye disease | atorvastatin | FDA-approved for other indications | Already used for this indication |
| vascular tumors of the eye | propranolol | FDA-approved for other indications | Already used for this indication |
| wet age-related macular degeneration | aflibercept | Already FDA-approved for this indication | Already used for this indication |
| wet age-related macular degeneration | bevacizumab | FDA-approved for other indications | Already used for this indication |
| wet age-related macular degeneration | omega-3 fatty acids | Available without FDA approval | Already used for this indication |
| wet age-related macular degeneration | pegaptanib sodium | Already FDA-approved for this indication | Already used for this indication |
| wet age-related macular degeneration | rapamycin (sirolimus) | FDA-approved for other indications | Cell-based/bench studies |

Of the 180 medications suggested by ChatGPT, 143 (79%) were FDA-approved for general medical use. However, only 32 out of 180 (18%) were specifically approved for the recommended ophthalmological conditions. Of all suggested medications, 41/180 (23%) were related to the retina, 82/180 (46%) were related to the cornea and anterior segment, 16/80 (8.9%) glaucoma, 22/180 (12%) Pediatrics and Strabismus, 1/180 (0.55%) neuro-ophthalmology, and 18/180 (10%) uveitis. Figure 1 summarizes the breakdown.

**Fig 1.**
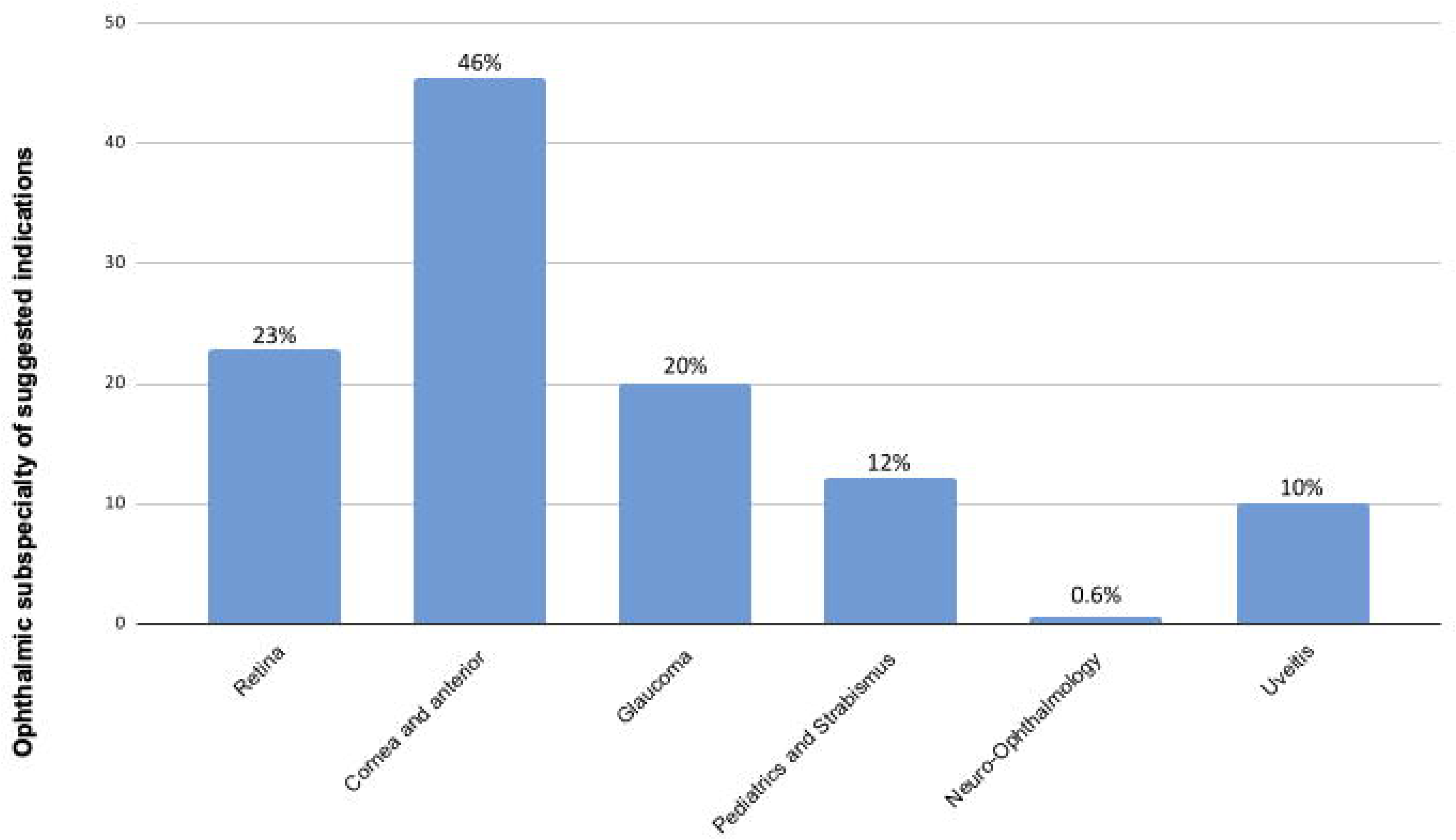
ChatGPT Medication Suggestions According to Ophthalmic Subspecialty. Breakdown of medications suggested by ChatGPT, categorized by ophthalmic subspecialties.

After conducting a comprehensive literature review to assess the level of evidence and novelty for these medications, it was determined whether they had been evaluated at the preclinical, clinical, or cell-based research levels, with the following findings: 2/180 (1.1%) were at the level of cell-based research, 15/180 (8.3%) have been tested in animal models, 86/180 (48%) were already used in the clinic, 30/180 (17%) were novel but supported by existing, limited literature, 27/180 (15%) repurposed for a novel ophthalmic use, 20/180 (11%) were novel for the disease but not supported by literature review and might be an unproductive suggestion by ChatGPT. Figure 2 summarizes these findings.

**Fig 2.**
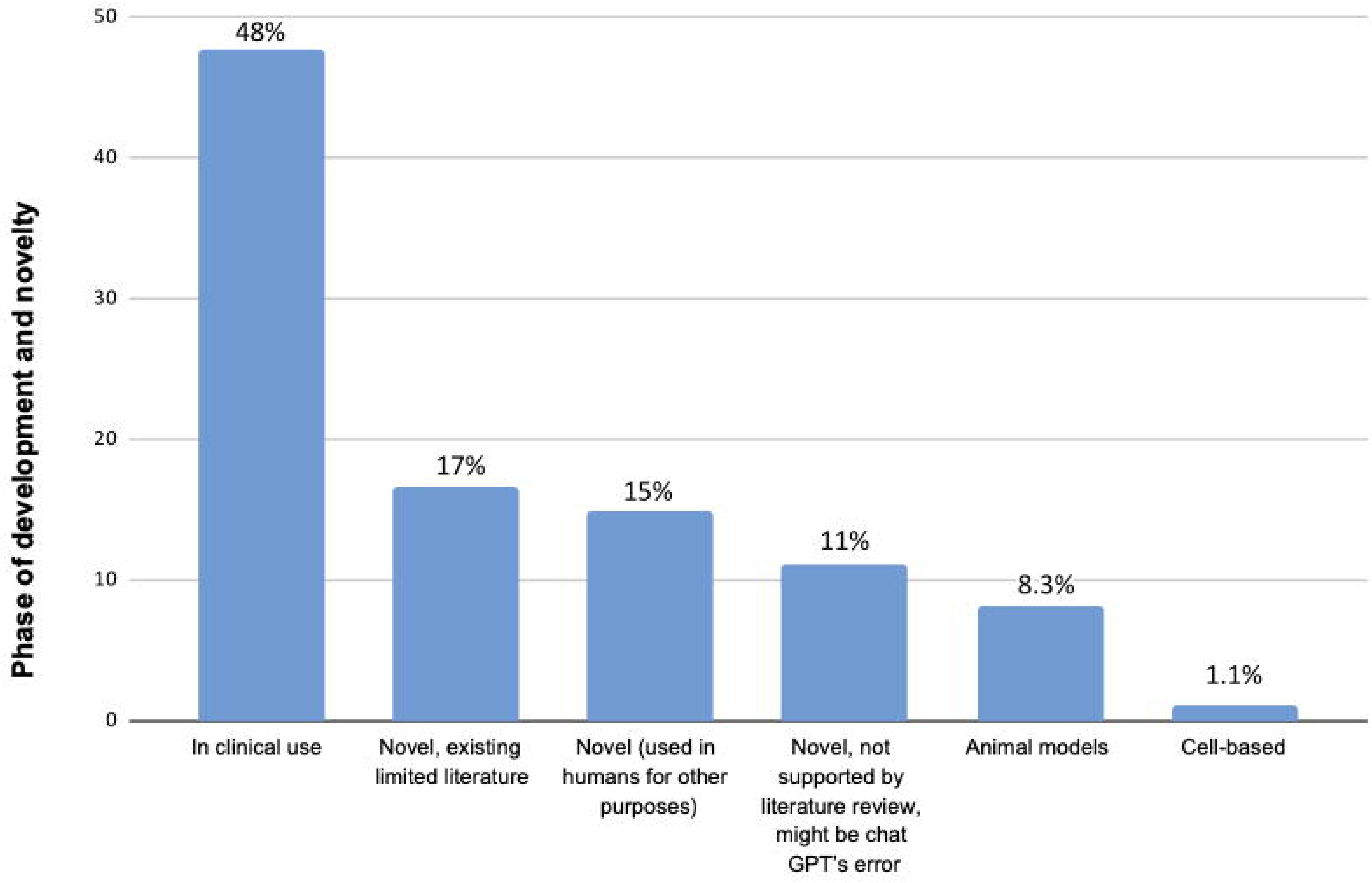
Level of Evidence and Novelty for Suggested Medications. Summary of the level of evidence and novelty for medications suggested by ChatGPT, categorized by research and clinical evaluation stages.

The level of novelty within each specialty is summarized in Figure 3. For instance, of 41 suggested treatments for retina-related diseases, 9 (22%) were at the level of animal models, 1 (2.4%) at the level of cell-based research, 10 (24%) had clinical applications, 5 (12%) likely unproductive suggestions, 10 (24%) novel and supported by existing, limited literature; and 6 (15%) were repurposed for a novel ophthalmic.

**Fig 3.**
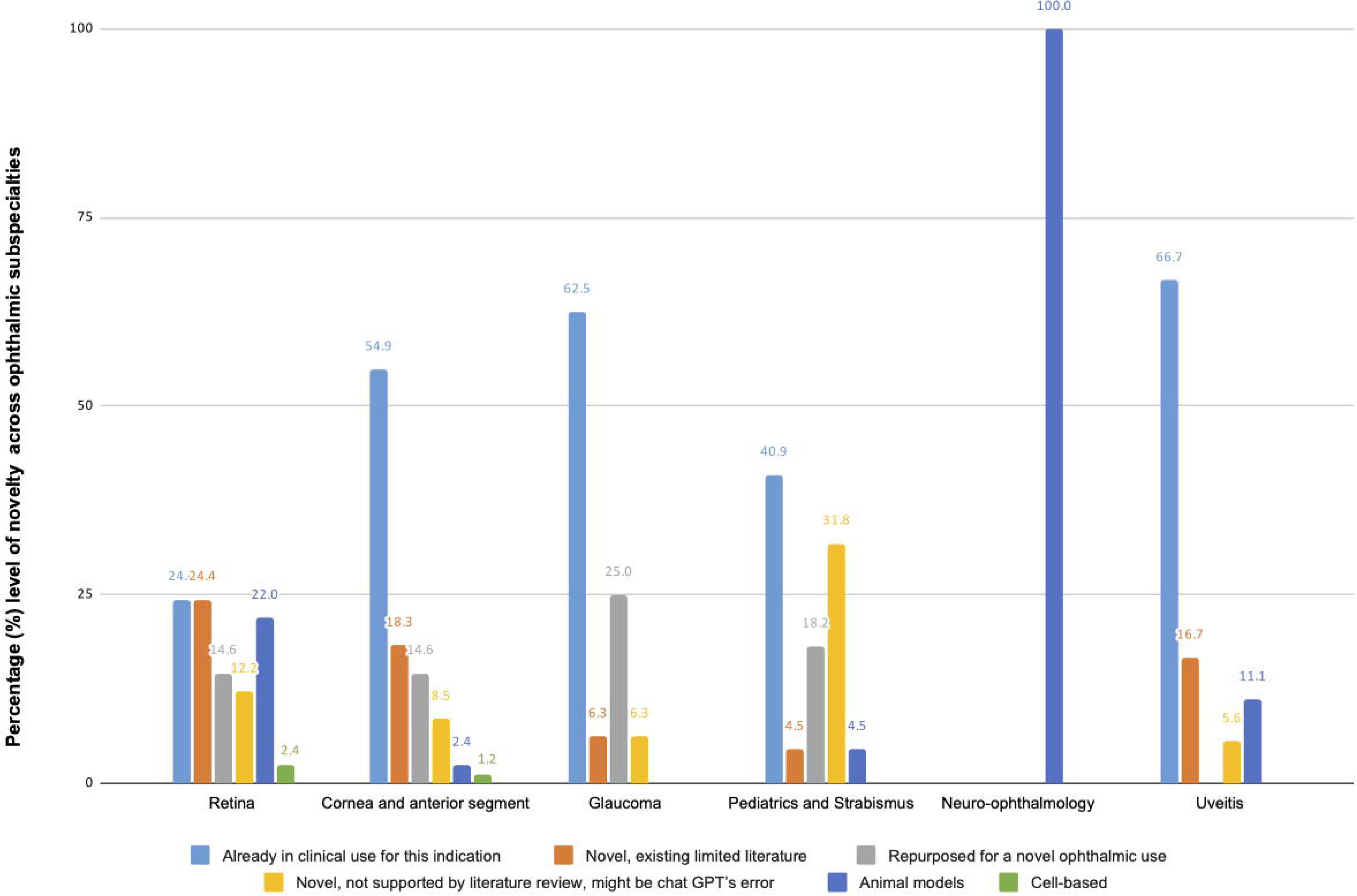
Level of Novelty for Suggested Medication Use Across Ophthalmic Subspecialties. Summary of the level of novelty for ChatGPT-suggested treatments across ophthalmic subspecialties.

## Discussion

This study evaluated 180 medications proposed by ChatGPT for 36 specific ophthalmic conditions. Among these, 79% were FDA-approved for general medical use, but only 18% were approved for the suggested ophthalmological indications. The recommendations covered various subspecialties, with the largest proportions targeting the cornea (46%) and retina (23%). A literature review showed that 47.8% were in clinical use, 16.7% were novel with limited supporting evidence, 15% were repurposed, and 11.1% lacked evidence.

While the suggestions provided by ChatGPT appear promising at first glance, most (48%) were for medications already widely used in clinical practice for specified purposes. For example, ChatGPT recommended moxifloxacin for bacterial conjunctivitis and brimonidine for glaucoma— both well-established treatments. Approximately 15% of the suggestions were novel repurposing of medications used in other fields, such as thalidomide for uveitis, sildenafil for retinal ischemia, or tizanidine for strabismus. These novel suggestions warrant further investigation to evaluate their safety and potential applications, offering new avenues for research in ophthalmology. However, 11% of the suggestions appeared to be unproductive suggestions, reflecting limitations in ChatGPT’s contextual understanding. These included recommendations such as acetazolamide for strabismus and tranexamic acid for preventing retinal detachment, likely resulting from keyword-based associations rather than a nuanced consideration of relevance or ChatGPT hallucinations [10]. The tendency of ChatGPT to “hallucinate” has been evidenced by its production of false scientific references and sources with improper DOI links [10]. This further emphasizes the crucial act of checking the accuracy of the information and sources provided by ChatGPT before their implementation. The remaining suggestions were either tested in animal models or based on limited case reports or series, yet were still categorized as novel treatments. We made an effort to critically evaluate these and distinguish them from truly novel suggestions.

In relation to the potential for novel discoveries with personalized patient care, Gonzales et al mention the potential of artificial intelligence to identify various drug-drug interactions and translate this for use in personalized medicine for patient care [11]. In addition, the potential of artificial intelligence for use in personalized medicine is being tested by utilizing an individual’s genetic makeup to form and adjust treatments in all fields of medicine [12]. For instance, the potential of ChatGPT has been explored for use in personalized obesity treatments, yet further research is still needed before recommendation to the public [13]. Personalized medicine will allow for innumerable novel treatment variations by ChatGPT, but the program has still been found to have limitations before getting to this point.

While abundant in the use of AI in ophthalmology, the data thus far has been limited in its use for discovering novel ophthalmic treatments. One study compared different models of artificial intelligence chatbots in their accuracy in planning retinal detachment surgeries, with ChatGPT being found to be the most precise and accurate in comparison to other models such as Google Gemini [14]. Deep learning models also have increasing potential for aid in drug development and analysis. For example, the deep learning model PIONEER is able to determine the three-dimensional interactions between proteins and their environment, providing invaluable insight toward drug discovery [15].

In regards to novel drug suggestions specific to ophthalmology, OphGLM is a new large language model (LLM) that performs ophthalmology-specific diagnoses and assessments using visual capabilities [6]. Ming et al suggest that the capabilities of LLM’s will allow researchers to identify novel drug targets in uveitis treatment, allowing for a more streamlined innovation of medications [7]. The study did not, however, evaluate the use of ChatGPT and/or other artificial intelligence models in generating these novel drug uses in the present.

Seth et. al. prompted ChatGPT for novel research ideas regarding oculoplastic surgical research advancements; unfortunately, the researchers did not find the suggestions to be novel and thus do not consider ChatGPT to be a central source for innovative ideas [16]. The “regenerate response” feature was used repeatedly by these researchers in the hope that ChatGPT would regenerate a novel idea.

However, there is hope in the ability of large language models to provide novel treatment options in the field of ophthalmology. In regards to ChatGPT’s knowledge of ophthalmology topics, one study found that the newest version of ChatGPT, GPT-4, was able to pass a practicing ophthalmology written exam, while other ChatGPT models were not [17]. In addition, one laboratory investigated ChatGPT’s performance on OpthoQuestions, a question bank typically used for board exam preparation, two years in a row. The study found that ChatGPT achieved a score of 46% in January of 2023 and the newer ChatGPT-4 later achieved 84% on the same question set [18]. This demonstrates the potential for newer models of ChatGPT to generate more accurate and reliable novel therapeutic suggestions. The field of ophthalmology is open to constant innovation and technological advancement, not only with the recent use of ChatGPT but also with other components of artificial intelligence such as using Deep Learning Systems based on fundus photographs [19].

The strengths of this study include the utilization of modern technology to generate novel medical treatments and the use of several researchers to perform quality checks. Because the use of LLM’s, such as ChatGPT, is still being explored for daily use and translation to the medical setting, research exploring these topics is essential to the future of medicine. With patient needs and treatments constantly changing, the opportunity to utilize AI as a tool in formulating novel treatment options for patients is essential. For instance, as demonstrated in Table 1, several drugs in this study have a novel suggestion by ChatGPT for use in ophthalmic disease, when it was only FDA-approved and used for a non-ophthalmic process. Of significance, it is estimated that 50% of patients do not properly adhere to their prescribed medications, with cost being one of the main factors [20]. This study demonstrates the potential for patients to utilize drugs for multiple disease processes, which can ultimately improve medication adherence and health outcomes. In addition, all information incorporated in this study was checked by at least two different researchers, with an intensive literature review to support drug classifications.

There are several limitations of this study. To start, the version of ChatGPT used in this study is the January 2024, ChatGPT-3.5, based on knowledge up to January 2022. Since this study was first conducted, there have since been improvements to the ChatGPT models. The newest release is GPT-4o, which has a knowledge cutoff of October 2023 and is deemed more capable overall, according to the official OpenAI Platform [21]. Thus, newer models of ChatGPT might have performed better. This study is worth repeating with the newest version of ChatGPT, as well as other AI platforms, to generate the most updated information. In addition, several of the results were repetitive or difficult to classify, with some recommendations even being deemed nonsensical. In fact, there have been findings of ChatGPT-3.5 providing hallucinated content, such as inaccurate title, author, or publication year, when asked to generate citations or references [22]. This further supports the need for human researchers, and physicians, to verify findings generated by AI.

## Conclusion

In summary, our study investigated the ability of ChatGPT to generate novel therapeutic recommendations in ophthalmology. The findings suggest that ChatGPT can formulate novel treatment options for a wide range of ophthalmic diseases. Of the suggestions, 27/180 (15%) appeared novel and reasonable, with promising potential for drug repurposing. These findings and options for drug repurposing should be evaluated with further laboratory and clinical research, as well as utilizing the most updated forms of AI alongside our methodology to further investigate additional potential novel treatment options.

## Data Availability

All data produced in the present work are contained in the manuscript.

## Acknowledgments

No acknowledgments

## References

1. Shen Y, Heacock L, Elias J, Hentel KD, Reig B, Shih G, et al. ChatGPT and Other Large Language Models Are Double-edged Swords. Radiology. 2023 Apr;307(2):e230163.

2. Abdullah YI, Schuman JS, Shabsigh R, Caplan A, Al-Aswad LA. Ethics of Artificial Intelligence in Medicine and Ophthalmology. Asia-Pac J Ophthalmol. 2021 May 1;10(3):289– 98.

3. Wang R, Feng H, Wei GW. ChatGPT in Drug Discovery: A Case Study on Anticocaine Addiction Drug Development with Chatbots. J Chem Inf Model. 2023 Nov 27;63(22):7189– 209.

4. Kozel G, Gurses ME, Gecici NN, Gökalp E, Bahadir S, Merenzon MA, et al. Chat-GPT on brain tumors: An examination of Artificial Intelligence/Machine Learning’s ability to provide diagnoses and treatment plans for example neuro-oncology cases. Clin Neurol Neurosurg. 2024 Apr 1;239:108238.

5. Filippi S. Measuring the Impact of ChatGPT on Fostering Concept Generation in Innovative Product Design. Electronics. 2023 Jan;12(16):3535.

6. Biswas S, Davies LN, Sheppard AL, Logan NS, Wolffsohn JS. Utility of artificial intelligence-based large language models in ophthalmic care. Ophthalmic Physiol Opt. 2024;44(3):641– 71.

7. Tan Yip Ming C, Rojas-Carabali W, Cifuentes-González C, Agrawal R, Thorne JE, Tugal-Tutkun I, et al. The Potential Role of Large Language Models in Uveitis Care: Perspectives After ChatGPT and Bard Launch. Ocul Immunol Inflamm. 2024 Aug 8;32(7):1435–9.

8. Huang AS, Hirabayashi K, Barna L, Parikh D, Pasquale LR. Assessment of a Large Language Model’s Responses to Questions and Cases About Glaucoma and Retina Management. JAMA Ophthalmol. 2024 Apr 1;142(4):371–5.

9. Drugs@FDA: FDA-Approved Drugs [Internet]. [cited 2025 Jan 2]. Available from: https://www.accessdata.fda.gov/scripts/cder/daf/index.cfm

10. Goddard J. Hallucinations in ChatGPT: A Cautionary Tale for Biomedical Researchers. Am J Med. 2023 Nov 1;136(11):1059–60.

11. Blanco-González A, Cabezón A, Seco-González A, Conde-Torres D, Antelo-Riveiro P, Piñeiro Á, et al. The Role of AI in Drug Discovery: Challenges, Opportunities, and Strategies. Pharmaceuticals. 2023 Jun 18;16(6):891.

12. Patrinos GP, Sarhangi N, Sarrami B, Khodayari N, Larijani B, Hasanzad M. Using ChatGPT to predict the future of personalized medicine. Pharmacogenomics J. 2023 Nov;23(6):178– 84.

13. Arslan S. Exploring the Potential of Chat GPT in Personalized Obesity Treatment. Ann Biomed Eng. 2023 Sep;51(9):1887–8.

14. Carlà MM, Gambini G, Baldascino A, Giannuzzi F, Boselli F, Crincoli E, et al. Exploring AI-chatbots’ capability to suggest surgical planning in ophthalmology: ChatGPT versus Google Gemini analysis of retinal detachment cases. Br J Ophthalmol. 2024 Sep 20;108(10):1457– 69.

15. Medscape [Internet]. 2024. The Protein Problem: The Unsolved Mystery of AI Drug Dev. Available from: https://www.medscape.com/viewarticle/protein-problem-unsolved-mystery-ai-drug-dev-2024a1000oje

16. Seth I, Bulloch G, Xie Y, Zhu Z. Exploring the Potential of ChatGPT for Advancing Oculoplastic Surgical Research. Annals of Ophthalmology. 2023;1038.

17. Lin JC, Younessi DN, Kurapati SS, Tang OY, Scott IU. Comparison of GPT-3.5, GPT-4, and human user performance on a practice ophthalmology written examination. Eye. 2023 Dec;37(17):3694–5.

18. Mihalache A, Huang RS, Popovic MM, Muni RH. Performance of an Upgraded Artificial Intelligence Chatbot for Ophthalmic Knowledge Assessment. JAMA Ophthalmol. 2023 Aug 1;141(8):798–800.

19. Li JPO, Liu H, Ting DSJ, Jeon S, Chan RVP, Kim JE, et al. Digital technology, tele-medicine and artificial intelligence in ophthalmology: A global perspective. Prog Retin Eye Res. 2021 May 1;82:100900.

20. Rohatgi KW, Humble S, McQueen A, Hunleth J, Chang SH, Herrick C, et al. Medication adherence and characteristics of patients who spend less on basic needs to afford medications. J Am Board Fam Med JABFM. 2021;34(3):561–70.

21. OpenAI Platform [Internet]. [cited 2025 Jan 2]. Available from: https://platform.openai.com

22. Chelli M, Descamps J, Lavoué V, Trojani C, Azar M, Deckert M, et al. Hallucination Rates and Reference Accuracy of ChatGPT and Bard for Systematic Reviews: Comparative Analysis. J Med Internet Res. 2024 May 22;26:e53164.

